# Forecasting the outbreak of COVID-19 in Lebanon

**DOI:** 10.1101/2020.09.03.20187880

**Authors:** Omar El Deeb, Maya Jalloul

## Abstract

This note explores the spread of the Coronavirus disease 2019 (COVID-19) in Lebanon using available data until August 25th, 2020 and forecasts the number of infections until the end of September using four diffierent scenarios for mitigation measures reflected in the reproductive number *R_t_*. Mitigation measures in Lebanon date back to early March soon after the first confirmed cases, and have been gradually lifted as of May. Thereafter, the country has witnessed a slow yet steady increase in the number of cases that has been significantly exacerbated after the explosion at Beirut harbor on August 4. Furthermore, we estimate the daily active cases in need of intensive care compared to the available number of beds and we assess accordingly that this capacity will be exhausted within a short span of time, unless severe measures are imposed.

## 1 Introduction

The COVID-19 outbreak in Lebanon has started on February 21, 2020 with few confirmed cases during the first days [1, 2]. Mitigation measures have been made into effiect early March, and have been gradually lifted as of May 10, while the airport activity was resumed with one-fifth capacity on July 1. A couple of weeks after, the number of infections was on the rise once again. On August 4, a massive explosion hit Beirut’s port caused by approximately 3000 tonnes of unsecured Ammonium Nitrate, leading to hundreds of deaths and thousands of injuries, in addition to a wide radius of destruction affiecting thousands of buildings, including hospitals, in and outside the city [3]. Hospitals and medical centers in Beirut and the neighboring regions experienced overflows in wounded patients, largely exceeding their capacities and resources, which is believed to have triggered a surge in the number of COVID-19 infections. The authorities decided subsequently to impose a partial lockdown, banning public gatherings, closing businesses and enforcing nighttime curfew starting August 21. A report issued by the Ministry of Public Health states that the available number of beds in the intensive care units that would be available to accommodate Coronavirus patients in all equipped hospitals stands at 336 beds [4], which establishes an upper bound for the capacity of the health sector in the fight against the virus.

## 2 Findings

We use the STEIR, a novel adaptation of the SEIR models accounting for trave, that was introduced in [5]. We employ data available until August 26, 2020 [1] to construct forecast scenarios of the spread of the disease until the end of September, 2020 (see Figure (1)).

**Figure 1:**
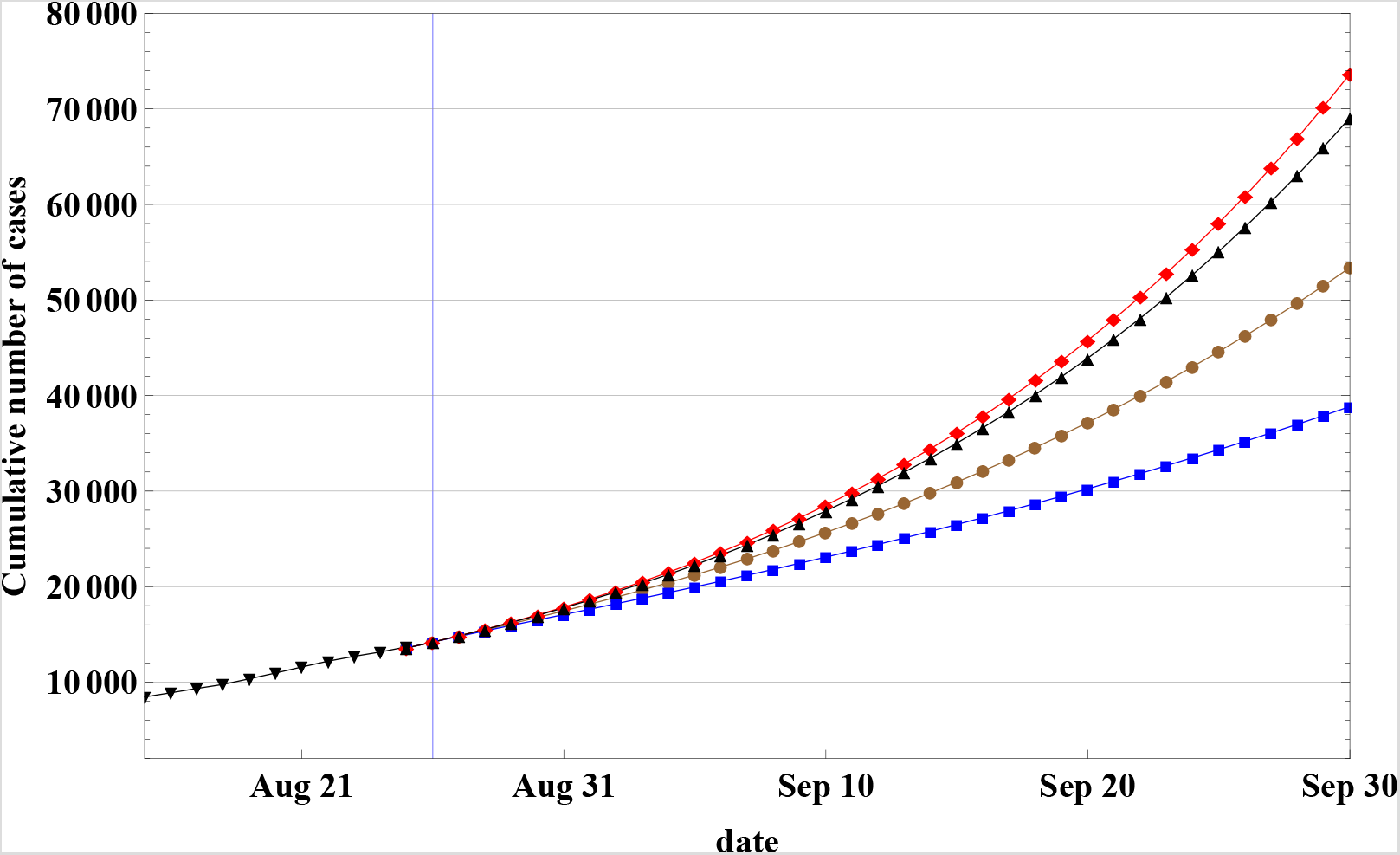
Cumulative number of infections according to the STEIR model in Lebanon, with appropriate parameterization of *R_t_*. The vertical blue line corresponds to day 186, which occurs on August 26, 2020. The black line before represents actually registered cases until that date, while the following four blue, brown, black and red lines correspond to future predictions based on four diffierent scenarios with *R_t_* = 1.5, 2, 2.4 or 2.5 depending on the extent of relaxation.

The model divides the population *N* into five categories of people: susceptible *S*, incoming travellers *T*, exposed *E*, infectious *I* and removed *R* (through recovery or death). The key parameters that determine the dynamics of spread are: the rate of exit by recovery or death per-day which is associated to the average illness period (*γ*), the rate at which exposed individuals become infected associated to the mean incubation period (*σ*), the reproductive number *R_t_* that determines the transmission rate from susceptible to infected and is a proxy for social distancing measures, the daily rate of incoming travellers (*τ*), and the average infection rate among the travellers (*θ*).

The numerical values of these parameters are consistent with those used in [5], apart from the parameterization of the basic reproduction number *R_t_* which we update according to the actual cases registered until August 26. We forecast four possible future scenarios for the development of *R_t_* corresponding to four levels of social distancing measures (see Figure 2), and we parameterize it as follows:

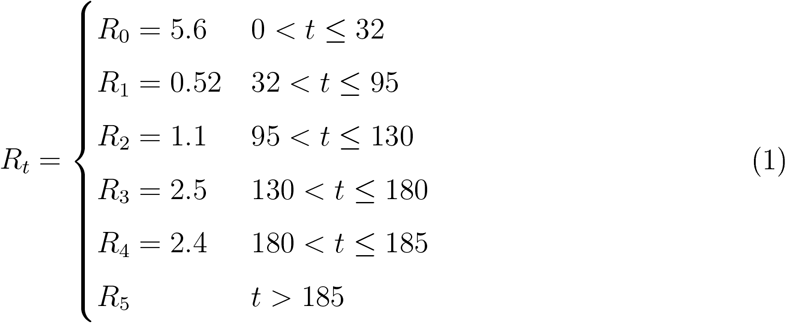

**Figure 2:**
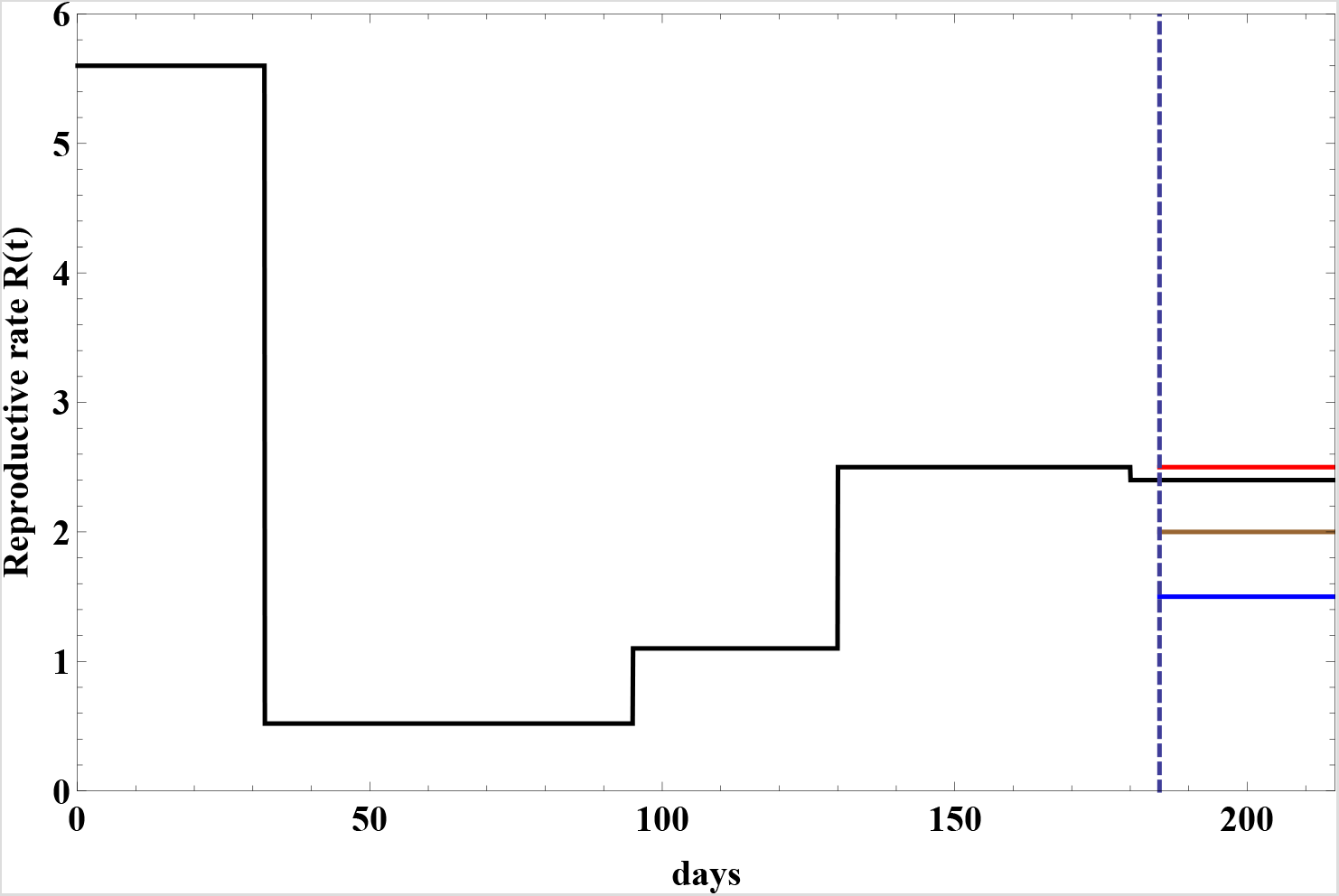
Change in the reproduction transmission factor *R*(*t*) as a function of time *t* (in days) in Lebanon. It starts at *R*_0_ = 5.6 then falls down at *t* = 32 days to *R*_1_ = 0.52 after strong mitigation measures. It rises to *R*_2_ = 1.1 at *t* = 95 days then to *R*_3_ = 2.5 at *t* = 130 days, before a slight decrease into *R*_4_ = 2.4 between *t* = 180 and *t* = 186. The four future possible scenarios inspected here have values of *R_t_* = *R*_5_ = 1.5, 2, 2.4 or 2.5 after *t* = 186 represented in blue, brown, black and red respectively.

where *R*_5_ is the future expected value for the reproduction number. Note that all values of *R*_0_ to *R*_4_ are obtained by fitting the reproduction rate according to the available data. We examine four potential values of the reproduction rate: *R*_5_ = 1.5 or 2 corresponding to intensifying the social distancing measures which could include complete lockdown, isolation of certain areas, imposing curfews, etc., 2.4 maintaining the current infection rate, and 2.5 which would lead to a slight increase with respect to the number of cases recorded in the last days. The corresponding scenarios of the spread are plotted in Figure(1) until the end of September.

In Figure (3), we plot the expected number of patients who would potentially need intensive care. This is determined from the forecast of the number of actively infected people predicted by the STEIR model, multiplied by the average rate of patients who need intensive care. According to publically available data in Lebanon [1], this rate stood at 0.98% in August.

**Figure 3:**
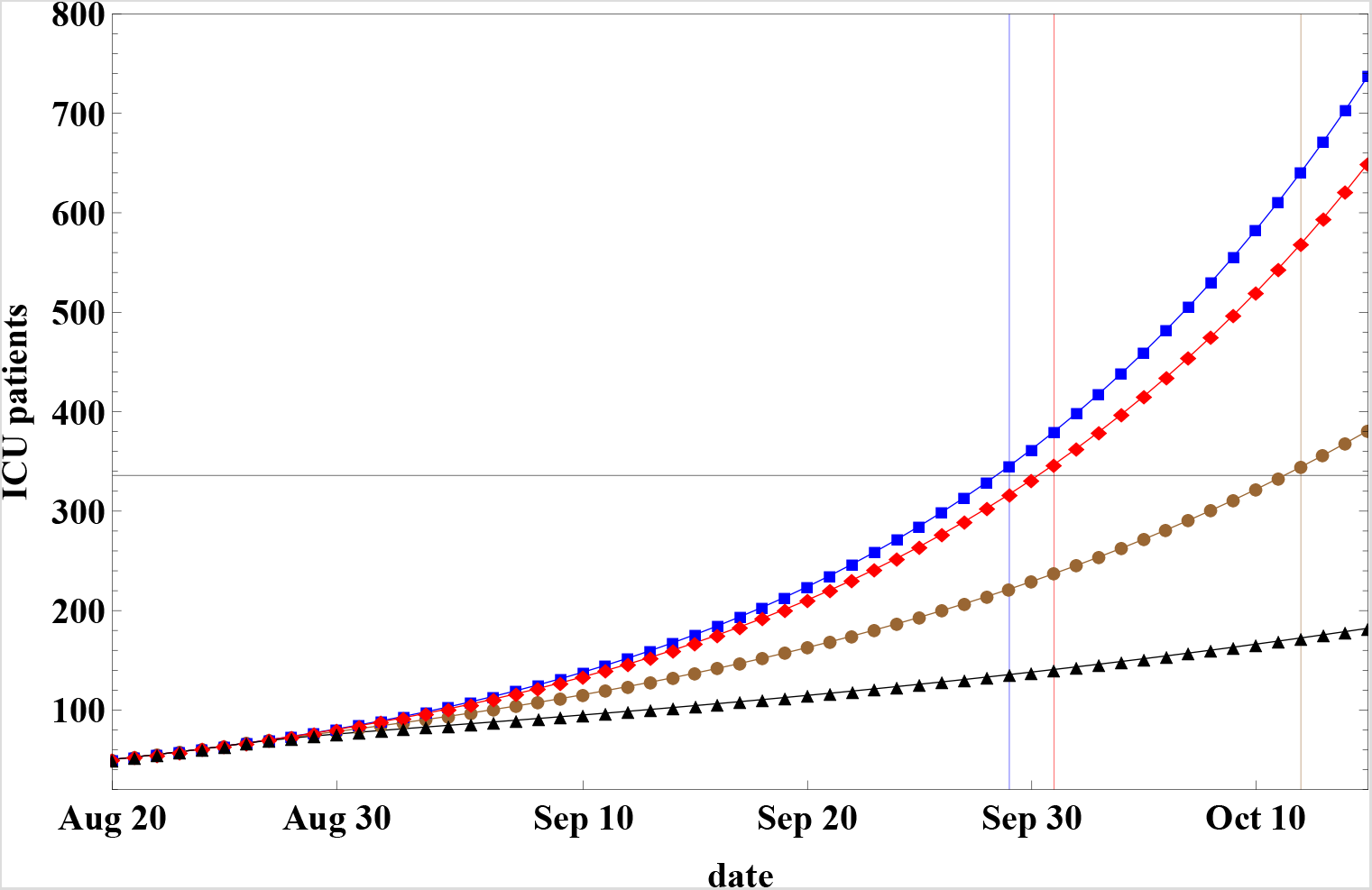
Expected number of ICU patients until the middle of October, according to the diffierent scenarios. The number of ICU patients would exceed the number of all available ICU beds (336 beds represented by the horizontal black line), on September 29, October 1, October 12 for *R*_5_ = 2.5, 2, 4 or 2 respectively. In case of severe measures with *R*_5_ = 1.5, the number of patients in need of ICU would be at nearly half the number of available beds by mid October.

## 3 Discussions and conclusions

Our results presented in Figure (1) and Table (1) reveal that the extent of the spread might reach up to 73655 cases by the end of September in case of relaxation of measures resulting in *R*_5_ = 2.5, in contrast to the case of severe measures corresponding to *R*_5_ = 1.5 which would generate about half of this number, amounting to 38839 cases. The number of daily infections on September 30 would reach 3421 cases in the relaxed scenario, nearly four times the 933 cases under the strict mitigation measures scenario.

**Table 1:** Expected numbers of daily and cumulative infections resulting from High (*R*_5_ = 1.5) and Low (*R*_5_ = 2.5) levels of mitigation measures on September 10, 20 and 30 respectively.

|  | Daily Infections |  | Cumulative number of Infections |  |
| --- | --- | --- | --- | --- |
|  | H | L | H | L |
| Sep 10 | 624 | 1192 | 21804 | 25946 |
| Sep 20 | 747 | 1929 | 28700 | 41635 |
| Sep 30 | 933 | 3421 | 38839 | 73655 |

The number of intensive care patients as displayed in Figure (3) would exhaust the capacity of the entire health sector by September 29 or October 1 in absence of strict mitigation measures. In the third scenario corresponding to *R*_5_ = 2, the maximum capacity would be attained by October 11. Imposing strict mitigation measures as in the fourth scenario, would shift the saturation level further until November.

Finally, it is essential to note that the estimated number of beds in the intensive care units used here and by the Ministry of Public Health is not exclusive for treatment of COVID-19 patients. Hence, actual saturation dates of the capacity of the health sector are likely to be earlier than the aforementioned. In this sense, it is of high importance to adopt the reasonable mitigation policies [6] to avoid bearing the consequences of a entirely overloaded health care system.

## Data Availability

Data available upon request.

